# Adjusting Coronavirus prevalence estimates for laboratory test kit error

**DOI:** 10.1101/2020.05.11.20098202

**Authors:** Christopher T. Sempos, Lu Tian

**Author notes:** These authors contributed equally to this article. Corresponding Author: Christopher T. Sempos, Ph.D.

## Abstract

Testing representative populations to determine the prevalence or percent of the population with active SARS-Cov-2 (COVID-19) infection and/or antibodies to infection is being recommended as essential for making public policy decisions to open-up or to continue enforcing national, state and local government rules to “shelter-in-place”. However, all laboratory tests are imperfect and have estimates of sensitivity and specificity less than 100% - in some cases considerably less than 100%. That error will lead to biased prevalence estimates. If the true prevalence of COVID-19 is low, possibly in the range of 1-5%, then testing error will lead to a constant background of bias that will most likely be larger and possibly much larger than the true prevalence itself. As a result, what is needed is a method for adjusting prevalence estimates for testing error. In this paper we outline methods for adjusting prevalence estimates for testing error both *prospectively* in studies being planned and *retrospectively* in studies that have been conducted. The methods if employed would also help to harmonize study results within countries and around the world. Adjustment can lead to more accurate prevalence estimates and to better policy decisions.

## Implications of Test Kit Error

Testing for the SARS2 Coronavirus (COVID-19) or antibodies to it in representative populations is being recommended as essential for making public policy decisions to open-up or to continue enforcing national, state and local government rules to “shelter-in-place”. Important objectives of testing are to estimate the prevalence or percent of the population currently infected with the COVID-19 virus and/or to estimate the prevalence who have developed antibodies to the COVID-19 virus after exposure, i.e. IgM and IgG (1,2). While cross-sectional studies are useful in estimating the current prevalence and trends in prevalence over time, it must be realized that all laboratory tests have measurement error.

Two key statistics used to characterize laboratory performance are sensitivity and specificity. Sensitivity is defined as the proportion of the population who have the disease and who test positive (a/a+c) (Table 1). Specificity, on the other hand, is defined as the proportion of the population who do not have the disease and who test negative (d/b+d) (3).

**Table 1:**
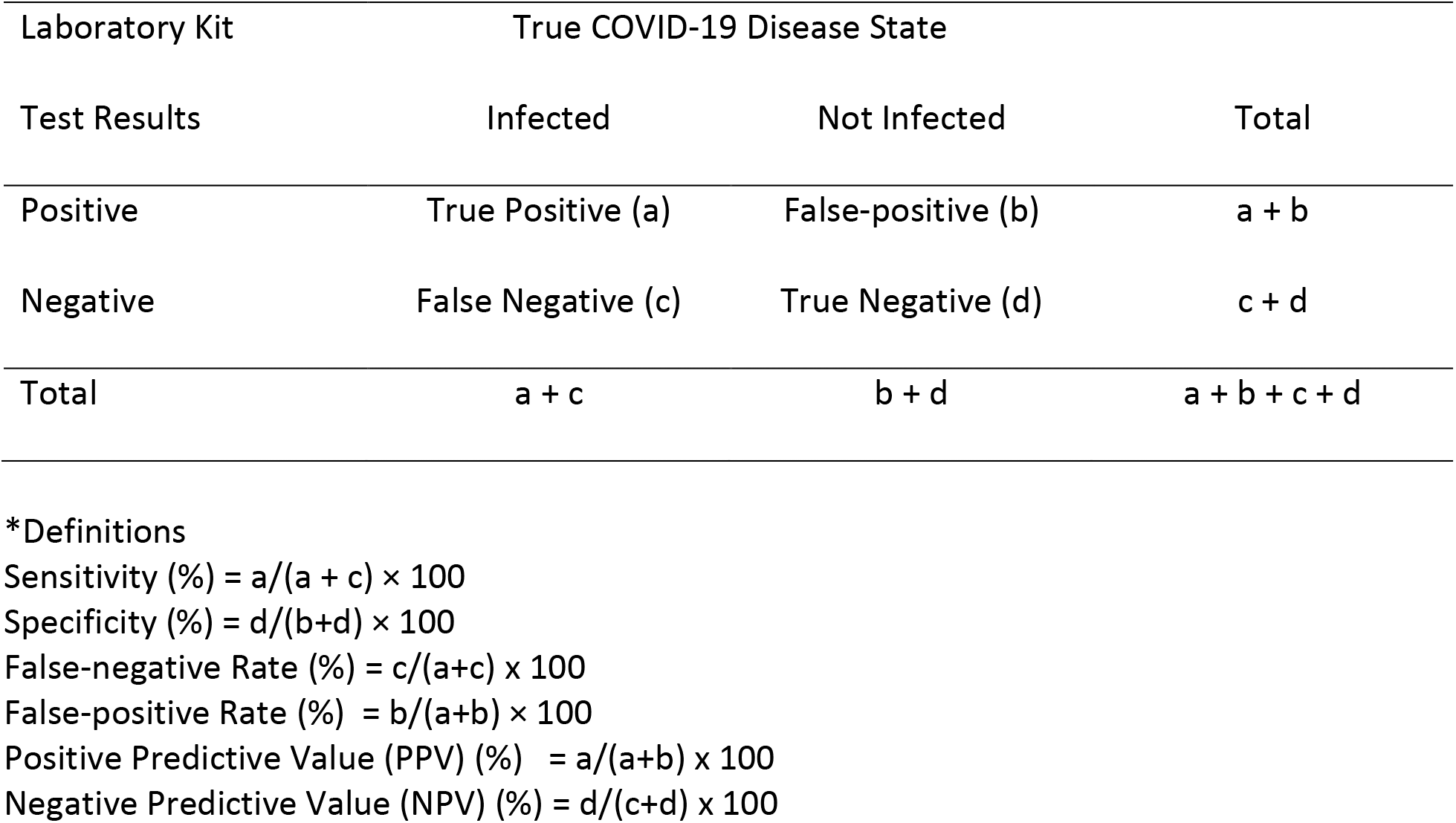
Theoretical Screening Table Used to Define Sensitivity, Specificity, and False-positive Rate*.

No laboratory test is 100% sensitive and specific and many will likely include substantial measurement error (4,5). That measurement error will result in biased prevalence estimates. Consequently, it is important to understand the impact of laboratory test error, how it changes with the “true prevalence” and to develop a strategy to correct or adjust for that error in estimating prevalence. In this paper, we will recommend a strategy to adjust prevalence estimates based on our experience, in successfully adjusting laboratory measurements of vitamin D as part of the Vitamin D Standardization Program (VDSP), and tailored to the unique circumstances surrounding COVID-19 testing (6,7).

To date most emphasis has been placed on the sensitivity of test kits to identify cases with COVID19 infection using, for example, reverse transcription polymerase chain reaction (RT-PCR) testing (8). That was done initially because the focus was on clinical diagnostic testing of people who displayed COVID-19 symptoms and/or at high risk of infections and the main concern was not to miss cases that should be quarantined in order to prevent the spread of the infection. Now, the focus of states and local governments is turning to documenting the percent of the population that has been infected with COVID-19 using immunoassays under the assumption that those individuals may have developed immunity that will last for some period of time in order to determine how and when to relax the “shelter-in-place” decrees.

However, the true Coronavirus prevalence estimate is currently thought to be quite low - possibly in the range of 1-5% - in many areas (9). In that case, it is essential to understand the impact of specificity in addition to sensitivity - as even small deviations of specificity from 100% may lead to identifying a set of positive samples which is largely composed of False-Positives.

For example, assume that a cross-sectional study is being conducted to determine the percentage of the population that is currently infected or has been infected by COVID-19. Moreover, let’s assume that the testing kits of interest have outstanding performance characteristics: sensitivity = 100% and specificity = 99% (Table 2). Let’s also assume that the true COVID-19 prevalence rate among those tested is 1%.

**Table 2:**
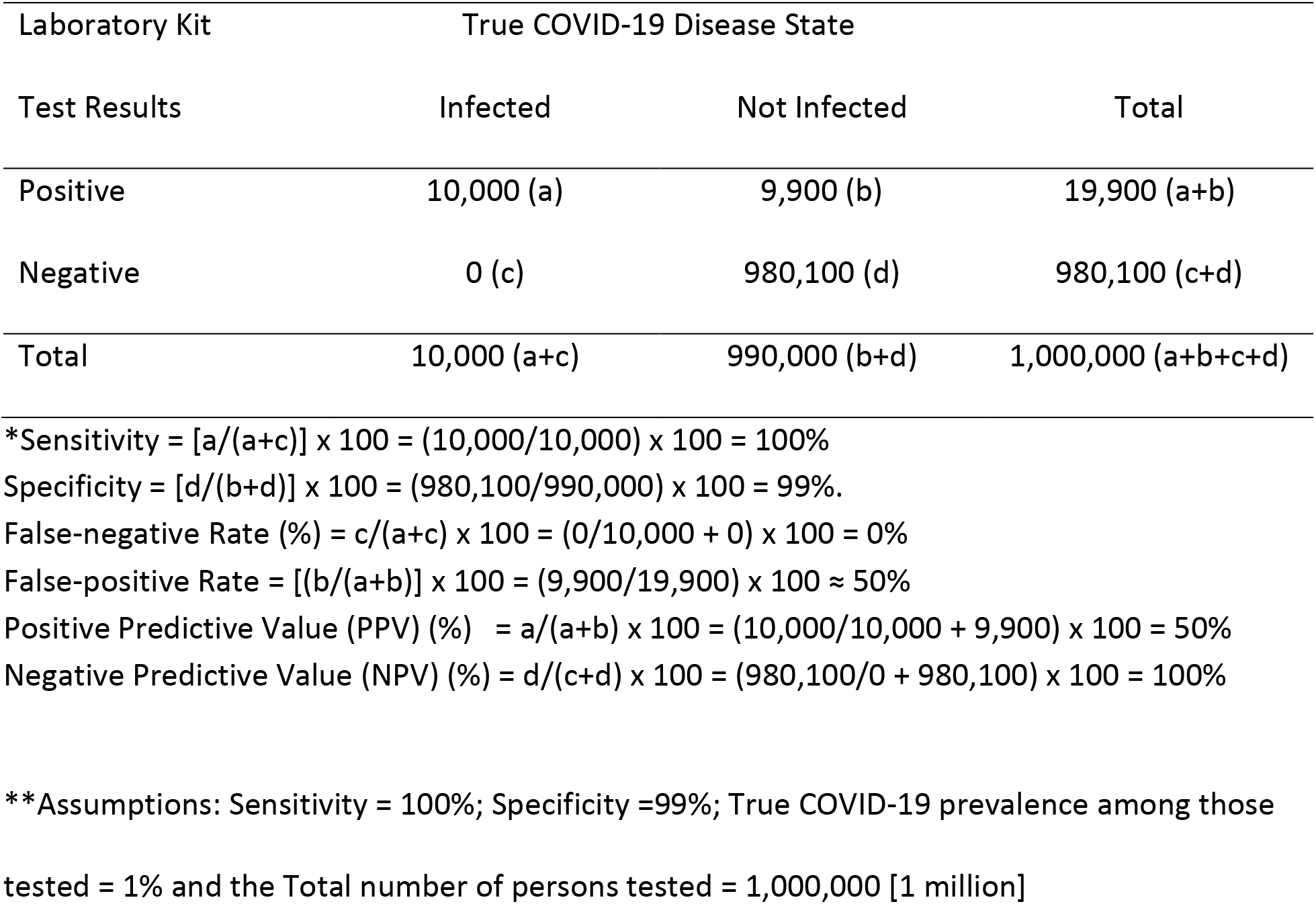
Screening Table Results* for Example Assumptions **

| Laboratory Kit<br>Test Results | True COVID-19 Disease State |  |  |
| --- | --- | --- | --- |
|  | Infected | Not Infected | Total |
| Positive | 10,000 (a) | 9,900 (b) | 19,900 (a+b) |
| Negative | 0 (c) | 980,100 (d) | 980,100 (c+d) |
| Total | 10,000 (a+c) | 990,000 (b+d) | 1,000,000 (a+b+c+d) |

\*Sensitivity = $[a/(a+c)] \times 100 = (10,000/10,000) \times 100 = 100\%$
Specificity = $[d/(b+d)] \times 100 = (980,100/990,000) \times 100 = 99\%$ .
False-negative Rate (%) = $c/(a+c) \times 100 = (0/10,000 + 0) \times 100 = 0\%$
False-positive Rate = $[(b/(a+b))] \times 100 = (9,900/19,900) \times 100 \approx 50\%$
Positive Predictive Value (PPV) (%) = $a/(a+b) \times 100 = (10,000/10,000 + 9,900) \times 100 = 50\%$
Negative Predictive Value (NPV) (%) = $d/(c+d) \times 100 = (980,100/0 + 980,100) \times 100 = 100\%$
\*\*Assumptions: Sensitivity = 100%; Specificity = 99%; True COVID-19 prevalence among those tested = 1% and the Total number of persons tested = 1,000,000 [1 million]

Then among 1 million persons tested, 10,000 COVID-19 cases will be correctly identified as True Positives (*a*) by the test kit and there are no False-Negatives (c) – a sensitivity of 100% (a/a+c)*100) (Tables 1and 2). Among the 990,000 truly uninfected individuals, there will be 9,900 False-positives (b) and 980,100 True Negatives (d) based on a Specificity = 99% (d/b+d)*100). Therefore, the False-Positive Rate - the proportion of not infected with COVID-19 among all those who tested positive (3) - will be equal to 50%, i.e. [*b/(a+b)* x 100= [9,900/9,900 + 10,000)] x 100 = 49.7%]. At a prevalence of 5%, the False-positive rate will be 17%.

If on the other hand, the sensitivity and specificity are both 95% then when the true prevalence is 1%, the False-Positive rate then will be 83.94% (Figure). As the true prevalence increases the false-positive rate will decrease. At a true prevalence of 5% the false-positive rate will still be 50%.

These calculations apply both to studies to determine the presence of the virus in an individual using PCR, and to studies to determine the development of antibodies in response to an infection using immunoassays, e.g. IgM and IgG.

The three factors essential for estimating the false-positive rate are: (1) Sensitivity and (2) Specificity of the testing kit and (3) the proportion of true COVID-19 cases among all those tested. Therefore, depending on performance characteristics of the test kits in use, as COVID-19 testing becomes more common in the US and in other countries this may lead to dramatically inflated COVID-19 prevalence estimates.

Therefore, studies to determine prevalence in representative samples need to have a plan imbedded in their study design to determine the sensitivity and specificity of the laboratory test kits used. Moreover, because the laboratory error will vary from study to study even if the same test kit is used it is essential that each study include a “harmonization” plan so that study results are comparable. Based on our experience in standardizing the measurement of serum total 25-hydroxyvitamin [25(OH)D] as part of the VDSP (6,7), we propose a general plan where all representative studies would be adjusted and harmonized to a common base - in a manner similar to age-adjusting mortality data. That plan includes methods for adjusting prospectively studies being planned and adjusting retrospectively studies that have been completed.

COVID-19 test kits, both PCR and antibody, are generally qualitative tests that provide Yes/No results which is different from the situation for serum 25(OH)D tests which provide a continuous quantitative result. However, even in this situation it is still possible to develop a framework which can be used to adjust crude prevalence estimates to the “true” prevalence (9,10). The framework would consist of: (1) selecting an established well-validated test, with documented sensitivity and specificity as close to 100%, as possible, to use as the reference-point assay or test kit; (2) Using that test kit to develop a series of “True Positive” and “True Negative” test samples; and (3) Using that set of test samples to estimate the sensitivity/ specificity or positive predictive value/negative predictive value of the study test kit. As we will mention later it may also be important to know the sensitivity and specificity of the reference-point assay or test kit.

This is similar to what assay manufacturers are required to do in their validation of test kits, but which is often carried out under ideal conditions. As described below, the framework for determining levels of sensitivity and specificity should resemble normal conditions of use, as much as possible, and take into account sources of error including those that occur in the pre-analytical phase (4).

Estimates of sensitivity and specificity could then be used to adjust the crude prevalence estimates from representative surveys using Appendix equation #1. Specifically, the adjusted prevalence is estimated using the following formula

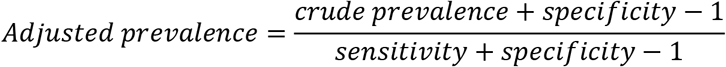

where the crude or observed prevalence is the proportion of the positive tests using the test kit, and sensitivity and specificity are their respective estimates. The entire adjustment process including calculating 95% confidence limits can be accomplished using, for example, EpiR software (10). Moreover, if everyone uses the same framework in every US state and in countries around the world, then data could be pooled to provide even larger datasets that could be used to study COVID19 in greater detail.

To summarize up to this point:

1. All test kits have measurement error, i.e. sensitivity and/or specificity < 100%.
2. Assay manufacturer estimates of sensitivity and specificity, are made under ideal conditions which may not reflect the true test kit sensitivity and specificity under actual field conditions.
3. Measurement error is the cumulative result of errors associated with biological sample collection, sample preparation, sample application to the test kit system and then the use of the test kit system to measure an individual biological sample for either the presence of the COVID-19 Virus or antibodies to it;
4. Laboratory tests tend to have a sensitivity or specificity near 100% but not generally both.
5. Numerous test kit systems for the measurement of the SARS-Cov-2 virus or antibodies to it will use a variety of different biological samples, e.g. nasal, nasopharyngeal and throat swabs, whole blood, and serum.

### Suggested Frameworks to Adjust for Test Kit Error

Two recent studies of COVID-19 antibody seroprevalence in Santa Clara County, California, USA and in Denmark suggest two general approaches for developing a framework to address those problems and needs (9,10). They are:

I. Select a Reference-point Assay to detect the presence of the COVID-19 Virus and/or one to detect IgM and IgG antibodies to the SARS-CoV_2 virus. The assays selected should be established and well validated assay, e.g. the WHO (11) or CDC RT-PCR assays (12) and the new CDC immunoassay (13). Those assays could then be the reference point in developing a set of True Positive and True Negative test samples as trueness controls that laboratories could use to determine the sensitivity and specificity of the test kits deployed in a prevalence study. This is the traditional approach.
II. The second approach includes the selection of Reference-point assays with a unique difference. Instead of using the Reference-point assays to develop a universal set of “Trueness Controls” that each study would use, each study would develop a set of “Positive” and “Negative” test samples using their measurement systems. Furthermore, those “Positive” and “Negative” samples from the study would then be sent to those with the reference point assay for verification. Verification by the Reference-point assay could then directly lead to an adjusted prevalence. The logic is as follows:

In the second approach, a set of test samples, collected, and prepared by as part of the harmonization framework of the prevalence study, will be sent to the “Reference-point” laboratory. The “Reference-point” test kit will be used to verify or validate the “Positive” and “Negative” test samples. Those results will be used to estimate the Positive Predictive Value (PPV) and Negative Predictive Value (NPV). The Positive Predictive Value (PPV) is the probability (P) that a “Positive” test sample is confirmed to be positive or a “case” using the Reference-point assay. The Negative Predictive Value (NPV) is the probability (P) that “Negative” test sample is confirmed to be negative or a “control” sample, again, using the Reference-point assay.

As shown in the Appendix, based on Option 2, we can then derive formulas for calculating the adjusted prevalence (P) based on equation #2 of the Appendix:

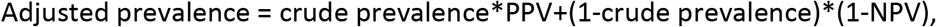

where PPV and NPV are their respective estimates.

Option I provides sensitivity and specificity estimates so that we can adjust the prevalence; while Option II provides PPV and NPV estimates, which can also be used to calculate the adjusted prevalence. In either case, the resulting adjusted prevalence is the same.

Another possible modification to Options 1 and 2 is to select <u>two reference-point assays.</u> One reference-point assay or test kit might have 100% sensitivity but unacceptable levels of specificity while another might be just the reverse. For example, an assay with 100% sensitivity could then be used to verify the studies “Positive” samples while the assay with 100% specificity would be used to verify the “Negative” study samples. As a result, using the two assays might then lead to a more precise prevalence estimate.

In all cases, it must be emphasized that in prevalence studies, as much as is possible, assay kit sensitivity and specificity “…should be evaluated for all the intended conditions of use…”, e.g. sample collection methods and lot to lot variation in test kits (4). As a result, Option II may provide the best possible approach for accomplishing that end.

Two further examples can help to show the potential impact of test kit error (14). For example, through April 28, 2020, 45,218 people in California tested positive for COVID-19 out of 526,084 tested. That is a crude or unadjusted prevalence of 8.6%. If all the tests used had a sensitivity of 95% and a specificity of 95%, then adjusted prevalence would be 4%, i.e. ([0.086 + 0.95 - 1]/[0.95 + 0.95 - 1])* 100 = 4% (Appendix) - less than half the crude prevalence of 8.6%. On the other hand, this combination of sensitivity and specificity corresponds to a PPV of 44.2% and an NPV of 99.8%. The adjusted prevalence using option II would again be 4%, i.e., [0.086*0.442+(1-0.998)×(1-0.086)]×100=4%.

For the second example, in New York State the number who tested positive [n=300,334] is a much higher percentage of all those tested [n=844,994; crude % = 36%] (14). Again, assuming that test kit sensitivity and specificity were both 95%, the adjusted percent is 34%. In this case adjustment, had little effect on the estimate of “true” prevalence.

These results reinforce the point discussed above and illustrated in the Figure, that when testing is restricted to symptomatic individuals, among which the true prevalence is high, the impact of test kit error is likely to be much less. But when testing is opened to all and especially in studies of representative samples where the true prevalence in many areas is likely to be small possibly on the order of 1-5%, adjustment for test kit error is essential in determining the true prevalence.

Using this or a similar set of guidelines would not only help to promote adjustment of prevalence estimates from representative studies around the world it would harmonize all results to one “standard”. That in-turn would guarantee comparability of study results from one locality to another in order to promote understanding of temporal and spatial trends in the COVID19 pandemic. That in turn, would promote the development of sound public policy.

Two final thoughts. First, assays have been and continue to be developed to measure antibody responses to the SARS-Cov-2 virus as a continuous variable (15). At this time, therefore, it may be useful for the field to begin discussing how those measurements can be standardize so that research/clinical results around the world are truly comparable. We believe that the methods developed by the VDSP for standardizing 25(OH)D measurement would be applicable and we suggest that they be taken up by this field, as well (7,16-18).

Second, to improve test kit accuracy assay manufacturers may wish to consider combining two assays into one single test kit. One component of the test kit would have nearly 100% sensitivity and the other would have nearly perfect specificity but the two tests together in one test kit would have nearly perfect sensitivity and specificity.

## Conclusions

All laboratory assays contain measurement error which needs to be estimated empirically. That is true of all COVID-19 assays. In representative cross-sectional COVID-19 studies to determine the proportion or prevalence of the population who is currently infected and/or the proportion or prevalence of the population with antibodies indicative of prior infection, even small deviations from 100% sensitivity and specificity will result in biased prevalence estimates. In this paper, we have outlined a series of steps than may be used to adjust representative studies for test kit error and to harmonize results over time and place in order to promote the development of effective public policy.

## Data Availability

Data in the paper are available for public sources.
No new data were generated in this research.

## Acknowledgments.

We would like to thank Drs. Peter Gergen, Karen W. Phinney and Dan S. Sharp for their kind help with thoughtful comments and suggestions for improving the paper.

---

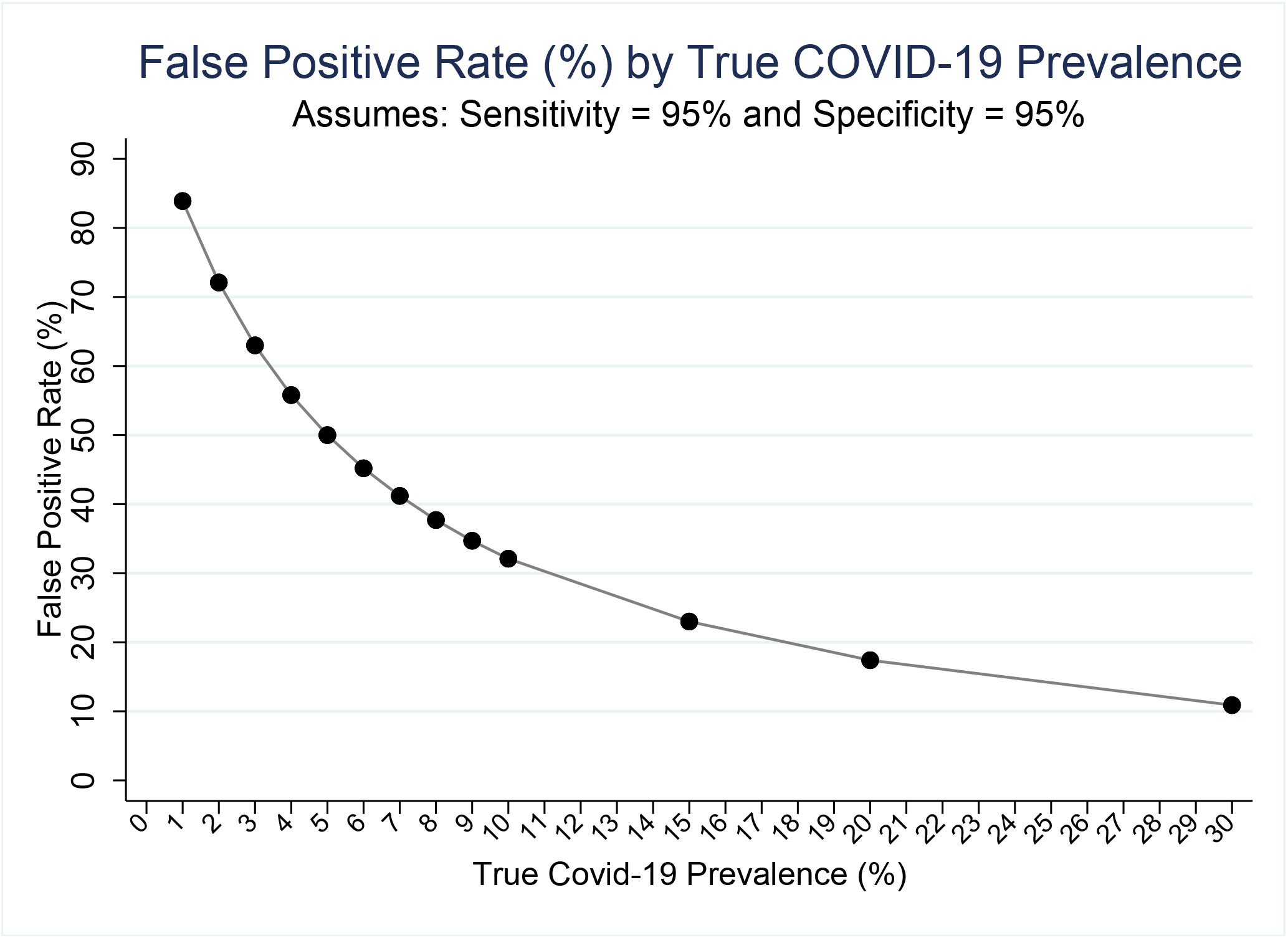

## Appendix

Deriving the adjusted prevalence estimate.

<u>In option I</u>, we observe the estimated sensitivity, specificity and

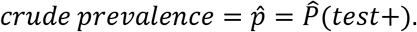

Noting the relationship

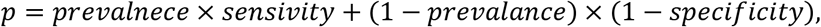

an adjusted prevalence estimate is

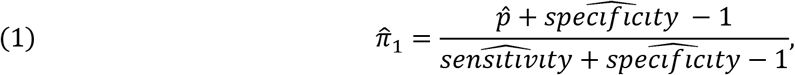

where 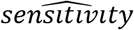 and 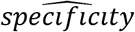 are estimated sensitivity and specificity based on the testing results on true positive and true negative samples, respectively. Furthermore, the false positive rate can be estimated by

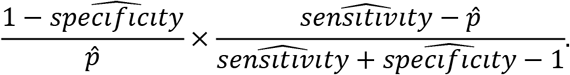

<u>In option II</u>, we observe the estimated *PPV = P(case*|*test+*), *NPV = P(control|test —)*, and

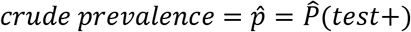

Based on the following system of equations:

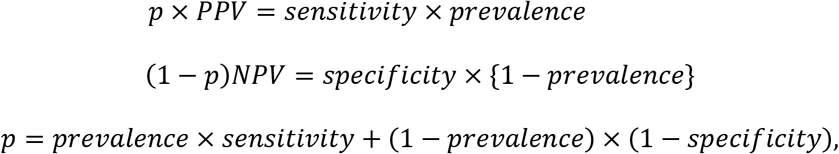

an adjusted prevalence estimate is

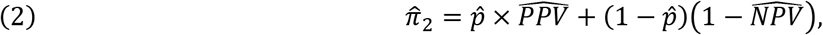

where 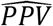 and 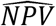 are estimated PPV and NPV based on the “reference-point” testing results on “positive” and “negative” samples from the study test, respectively. As a byproduct, we also can estimate the sensitivity and specificity of the test as

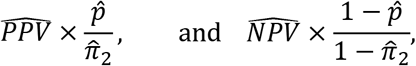

respectively.

## References

1. Beeching NJ, Fletcher TE, Beadsworth MBJ. Covid-19: testing times. BMJ. 2020 Apr 8;369:m1403. doi: 10.1136/bmj.m1403.

2. Sheridan C. Coronavirus and the race to distribute reliable diagnostics. Nat Biotechnol. 2020 Apr;38(4):382–384. doi: 10.1038/d41587-020-00002-2.

3. McCarthy N, Smith A. Chapter 2. Clinical Epidemiology. In: Abubakar I, Stagg HR, Cohen T, Rodrigues LC (eds). Infectious Disease Epidemiology. Oxford University Press. 2016.

4. Working document of Commission services. Current performance of COVD-19 test methods and devices and proposed performance criteria. EU Science Hub. https://ec.europa.eu/jrc/en/news/coronavirus-commission-issues-guidelines-testing

5. Whitman JD, Hiatt J, Mowery CT, Shy BR, et al. Test performance evaluation of SARS-CoV-2 serological assays. https://covidtestingproject.org/downloaded April 26, 2020.

6. Cashman KD, Holvik K, Andersen R, Linneberg A, Lamberg-Allardt C, Tetens I, Gronberg IM, Meyer HE, Keily M, Dowling K, Skrabakova Z, Lundqvist A, Koponen P, Sundvall J, Koskinen S, Nystrup LL, Thuesen BH, Sempos CT, Durazo-Arvizu RA, Skrakova Z. Standardizing serum 25-hydroxyvitamin D data from four Nordic population samples using the Vitamin D Standardization Program protocols: shedding new light on vitamin D status in Nordic individuals. Scand J Clin Lab Inves. 2015 75(7):549–561.

7. Sempos CT, Heijboer AC, Bikle DD, Bollerslev J; Bouillon R, Brannon PM, DeLuca HF, Jones G, Munns CF, Bilezikian JP, Giustina A, Binkley N. Vitamin D Assays and the Definition of Hypovitaminosis D: Results from the 1^st^ International Conference on Controversies in Vitamin D. Br J Cl Pharm. 2018;84(10):2194–2207. doi: 10.1111/bcp.13652.

8. West CP, Montori VM, Sampathkumar P, COVID-19 Testing: The Threat of False-Negative Results, Mayo Clinic Proceedings (2020), doi: https://doi.org/10.10167j.mayocp.2020.04.004.

9. Bendavid E, Mulaney B, Sood N, Shah S, Ling E, Bromley-Dulfano R, Lai C, Weissberg Z, Saavedra-Walker R, Tedrow J, Tversky D, Bogan A, Kupiec T, Eichner D, Gupta R, Ioannidis JPA, Bhattacharya J. COVID-19 Antibody Seroprevalence in Santa Clara County, California. medRxiv preprint doi: https://doi.org/10.1101/2020.04.14.20062463. Version 2, April 27, 2020.

10. Erikstrup C, Hother CE, Pedersen OBV, Mølbak K, Skov RL, Holm DK, Sækmose S, Nilsson AC, Brooks PT, Boldsen JK, Mikkelsen C, Gybel-Brask M, Sørensen E, Manh Dinh KM, Mikkelsen S, Møller BK, Haunstrup T, Harritshøj L, Jensen BA, Hjalgrim H, Lillevang ST, Ullum H. Estimation of SARS-CoV-2 infection fatality rate by real-time antibody screening of blood donors. medRxiv preprint doi: https://doi.org/10.1101/2020.04.24.20075291.this version posted April 28, 2020.

11. Corman VM, Landt O, Kaiser M, Molenkamp R, Meijer A, Chu DKW, Bleicker T, Brünink S, Schneider J, Schmidt ML, Mulders DGJC, Haagmans BL, van der Veer B, van den Brink S, Wijsman L, Goderski G, Romette JL, Ellis J, Zambon M, Peiris M, Goossens H, Reusken C, Koopmans MPG, Drosten C. Detection of 2019 novel coronavirus (2019-nCoV) by real-time RT-PCR. Euro Surveill. 2020 Jan;25(3). doi: 10.2807/1560-7917.ES.2020.25.3.2000045.

12. Centers for Disease Control and Prevention. Coronavirus Disease 2019 (COVID-19). CDC Viral Test for COVID-19. https://www.cdc.gov/coronavirus/2019-ncov/php/testing.html (Accessed April 30, 2020).

13. Freeman B, Lester S, Mills, Rasheed MAU, Moye S, Abiona O, Hutchinson GB, Morales-Betoulle M, Krapinunaya I, Gibbons, Chiang C-F, Cannon D, Klena J, Johnson JA, Owen SM, Graham BS, Corbett KS, Thornberg NJ. Validation of a SARS-CoV-2 spike protein ELISA for use in contact investigations and serosurveillance. bioRxiv preprint doi: https://doi.org/10.1101/2020.04.24.057323. This version posted April 25, 2020.

14. Worldometer. Coronavirus. https://www.worldometers.info/coronavirus/country/us/. (Accessed April 28, 2020).

15. Long QX, Liu BZ, Deng HJ, Wu GC, Deng K, Chen YK, Liao P, Qiu JF, Lin Y, Cai XF, Wang DQ, Hu Y, Ren JH, Tang N, Xu YY, Yu LH, Mo Z, Gong F, Zhang XL, Tian WG, Hu L, Zhang XX, Xiang JL, Du HX, Liu HW, Lang CH, Luo XH, Wu SB, Cui XP, Zhou Z, Zhu MM, Wang J, Xue CJ, Li XF, Wang L, Li ZJ, Wang K, Niu CC, Yang QJ, Tang XJ, Zhang Y, Liu XM, Li JJ, Zhang DC, Zhang F, Liu P, Yuan J, Li Q, Hu JL, Chen J, Huang AL. Antibody responses to SAS-Cov-2 in patients with COVID-19. Nat Med. 2020 Apr 29. doi: 10.1038/s41591-020-0897-1.

16. Tian L, Durazo-Arvizu RA, Myers G, Brooks S, Sarafin K, Sempos CT. The estimation of calibration equations for variables with heteroscedastic measurement error. Stat Med 2014;33:4420–4436.

17. Sempos C, Betz J, Camara J, Carter G, Cavalier E, Clarke M, Dowling K, Durazo-Arvizu R, Hoofnagle A, Liu A, Phinney K, Sarafin K, Wise S, Coates P. General Steps to Standardize the Laboratory Measurement of Serum Total 25-Hydroxyvitamin D. J AOAC Int. 2017; 100(5):1230–1233.

18. Durazo-Arvizu R, Tian L, Brooks SPJ, Sarafin K, Cashman KD, Kiely M, Merkel J, Myers GL, Coates PM, Sempos CT. The Vitamin D Standardization Program (VDSP) Manual for Retrospective Laboratory Standardization of Serum 25-Hydroxyvitamin D Data. J AOAC Int. 2017 100(5):1234–1243.

